# *Plasmodium vivax* genomic surveillance in the Peruvian Amazon with Pv AmpliSeq assay

**DOI:** 10.1101/2023.12.22.23300425

**Authors:** Johanna Helena Kattenberg, Luis Cabrera-Sosa, Erick Figueroa-Idelfonso, Mathijs Mutsaers, Pieter Monsieurs, Pieter Guetens, Berónica Infante, Christopher Delgado-Ratto, Dionicia Gamboa, Anna Rosanas-Urgell

## Abstract

**Background:** *Plasmodium vivax* is the most predominant malaria species in Latin America, constituting 71.5% of malaria cases in 2021. With several countries aiming for malaria elimination, it is crucial to prioritize effectiveness of national control programs by optimizing the utilization of available resources and strategically implementing necessary changes. To support this, there is a need for innovative approaches such as genomic surveillance tools that can investigate changes in transmission intensity, imported cases and sources of reintroduction, and can detect molecular markers associated with drug resistance.

**Methodology/Principal Findings:** Here, we apply a modified highly-multiplexed deep sequencing assay: Pv AmpliSeq v2 Peru. The tool targets a newly developed 41-SNP Peru barcode for parasite population analysis within Peru, the 33-SNP vivaxGEN-geo panel for country-level classification, and 11 putative drug resistance genes. It was applied to 230 samples from the Peruvian Amazon (2007 – 2020), generating baseline surveillance data. We observed a heterogenous *P. vivax* population with high diversity and gene flow in peri-urban areas of Maynas province (Loreto region) with a temporal drift. In comparison, in an indigenous isolated area, the parasite population was genetically differentiated (F_ST_=0.07-0.09) with moderate diversity and high relatedness between isolates in the community. In a remote border community, a clonal *P. vivax* cluster was identified, with distinct haplotypes in drug resistant genes and *ama1*, more similar to Brazilian isolates, likely representing an introduction of *P. vivax* from Brazil at that time. To test its applicability for Latin America, we evaluated the SNP Peru barcode in *P. vivax* genomes from the region and demonstrated the capacity to capture local population clustering at within-country level.

**Conclusions/Significance:** Together this data shows that *P. vivax* transmission is heterogeneous in different settings within the Peruvian Amazon. Genetic analysis is a key component for regional malaria control, offering valuable insights that should be incorporated into routine surveillance.

**Author Summary:** Latin America is aiming towards malaria elimination. Genomic surveillance is crucial for a country’s malaria strategy, aiding in understanding and stopping the spread of the disease. While widely used for another malaria species (*Plasmodium falciparum*), limited tools exist for tracking *P. vivax*, a significant player in malaria-endemic areas outside of Africa, and the primary cause of malaria in Latin America.

In this study, we used a new tool, Pv AmpliSeq v2 Peru assay, to examine the genetic makeup of malaria parasites in the Peruvian Amazon. This tool helps us see how the parasites from different areas are connected and tracks markers that could indicate resistance to drugs. We found that the parasites from remote areas in the Amazon were genetically different from parasites in areas surrounding the main city of Iquitos, and parasites in a remote border community were genetically more similar to Brazilian parasites.

We also show that the Pv AmpliSeq v2 Peru assay can be used to study parasites from other countries in Latin America, highlighting the broader application in the region. Considering that parasites are not constrained by borders and can easily spread between neighboring countries, a regional approach can be crucial for malaria elimination.

## Introduction

Malaria continues to impact millions of people around the world, particularly those living in low- and middle-income countries. *P. vivax* infections make up 18.0% to 71.5% of cases in regions outside Africa, with the highest proportions in the Americas [1], mainly affecting populations living in remote areas with poor access to healthcare services. The Amazon Basin reports the majority of malaria cases in the Americas, including Peru (> 26000 malaria cases in 2022, 88% caused by *P. vivax*) [2].

Challenges to malaria control and elimination in the Amazon region are diverse. *P. vivax* infections in the region are often asymptomatic (58-93%), below the detection threshold of microscopy (61-96%, sub-patent infections) [3], while a significant proportion of these infections carry gametocytes, the parasite stages responsible for transmission to the vector [4]. Moreover, the highest *P. vivax* incidence occurs in hard-to-reach areas, such as remote indigenous communities [5], and cross-border regions [6] where human mobility, often linked to economic activities, facilitates transmission and reintroduction [7]. In addition, low density infections typically require PCR-based tools for diagnosis, which are difficult to implement in remote areas. Although isothermal amplification assays -such as loop-mediated isothermal amplification (LAMP)- have the potential for point-of-care diagnosis [8–10], they have not been broadly implemented. Therefore, these asymptomatic and sub-patent infections typically are undiagnosed and untreated, maintaining malaria transmission [4].

Despite these *P. vivax* specific challenges, the region has made significant efforts to control malaria, with a reduction of the number of cases by 60% from 2000 to 2021 [1]. Therefore, many countries are moving towards malaria elimination in the next 5-10 years by implementing National Malaria Control Programs (NMCPs). In Peru, the Ministry of Health (MINSA) launched a National Malaria Elimination Program (NMEP) in 2022, intending to reduce the number of cases by 90% in 2030 [11].

*P. vivax* transmission is not uniform across the Peruvian Amazon region and varies significantly between different ecological niches suggesting that transmission is also influenced by ecological and environmental factors such as temperature and rainfall [12]. Since efforts to control *P. vivax* malaria in Peru must consider its complexity and heterogeneity of transmission, well-structured and integrated genomic surveillance systems can significantly contribute to pinpoint priorities and guide NMCP/NMEPs decision-making [13–15]. These systems empower countries to detect areas with the greatest need for control measures, identify program deficiencies and suboptimal strategies, and determine the amount of connectivity between regions. Through genomic analyses, the *P. vivax* parasite population of Latin America has been described as a distinct population from the rest of the world, presenting relatively lower genetic diversity due to relatively recent introductions in the region [16–18]. In the case of the Peruvian Amazon, Iquitos city acts as a source from where parasites spread to other areas through human mobility related to socio-economic activities [19, 20]. Additionally, there is substantial gene flow between parasite populations from geographically distant areas in the region [21].

Previously, we have developed a highly-multiplexed amplicon sequencing (AmpliSeq) assay for malaria genomic surveillance of *P. falciparum* in Peru (Pf AmpliSeq Peru, [22]) and *P. vivax* in Vietnam (Pv AmpliSeq v1 Vietnam, [23]). Our assays include validated and candidate genes associated with antimalarial resistance, SNP-barcodes to serve different functions, *i.e.* a global *P. vivax* SNP-barcode for prediction of origin, and country or region-specific SNP-barcodes for population genetics analysis such as transmission dynamics and connectivity. The AmpliSeq assays have the advantage, in addition to cover a large panel of genetic variants of interest, to be easily adaptable for diverse epidemiological contexts, and have been successfully used to identify temporal changes in the parasite population (before and after NMCP interventions), monitor drug resistant markers, assess connectivity, and identify sources of reintroduction.

In this study, we modified the Pv AmpliSeq v1 Vietnam to Pv AmpliSeq v2 Peru, incorporating a Peru-specific SNP-barcode with in-country resolution instead of the Vietnam barcode. To generate genetic data that can be used as baseline information for molecular surveillance in Peru, we used this assay to analyze *P. vivax* samples (N = 230) collected between 2007 and 2020 across 11 districts in the Peruvian Amazon. Finally, the Peru barcode was evaluated in a published dataset of 399 genomes from Latin America, achieving sufficient resolution to detect the previously reported population differentiation between countries. This tool provides invaluable information that can help in effectively guiding malaria control and elimination efforts in Peru by facilitating the generation of high-quality data and strategic information and can be used for malaria surveillance in Latin America.

## Methods

### SNP selection for barcode design

To design a SNP barcode with in-country resolution in Peru, raw whole genome sequencing data (Fast Q files) of *P. vivax* isolates from Peru generated in-house (n=30 from [23, 24]) was combined with online available Peruvian *P. vivax* genomes (n=100 from [25–28]) and jointly genotyped after variant calling as described elsewhere [17]. Briefly, FASTQ files were aligned to the PvP01 reference genome version 46 from PlasmoDB [29] and variants were called using GATK4 HaplotypeCaller. Allele frequencies of the selected SNPs in the design were assessed in the global genome dataset (n=1474) (17), as well as in genomes from Latin America (n=399) from that dataset.

To design the SNP Peru barcode, unlinked biallelic SNPs were filtered from the genomic dataset by LD-pruning in 5-6 iterations by scanning over the genome in 500 bp windows to remove uninformative SNPs with pairwise LD>0.2 using the python package scikit-allel. Subsequently, the contributions of the SNPs to genetic clusters were determined using discriminant analysis of principal components (DAPC) [30] with K-means inferred populations (n = 10) using the adegenet package in R. DAPC was performed multiple times (n=20) with cross-validation, and associated allele loadings between simulations were compared to determine the most contributing alleles. Finally, 49 SNPs with high allele loadings in the DAPC were selected with spread over the chromosomes (2-4 SNPs per chromosome) and distance (>500 bp) from drug resistant amplicons targeted in the assay to avoid linkage. The Illumina Concierge team (Illumina, San Diego, USA) used DesignStudio software with the PvP01 reference genome to add amplicons for the new barcode to the existing Pv AmpliSeq v1 Vietnam custom panel design [23] without the Vietnam SNP-barcode. Out of the 49 targeted SNPs, amplicon design was successful for 41 SNPs, resulting in the Pv Peru Barcode.

### Samples and study settings

In order to generate baseline genetic surveillance data that includes parasite populations from peri-urban areas, remote areas and border communities across a wide area in the Peruvian Amazon, we selected samples retrospectively from prior studies in Peru. *P. vivax* qPCR-positive samples (n=230) were selected based on geographical origin and parasite density (≥5 parasites/µL by qPCR). Samples were from 11 districts in 3 provinces in the Loreto region: Loreto, Maynas, and Mariscal Ramon Castilla (Fig 1).

**Figure 1.**
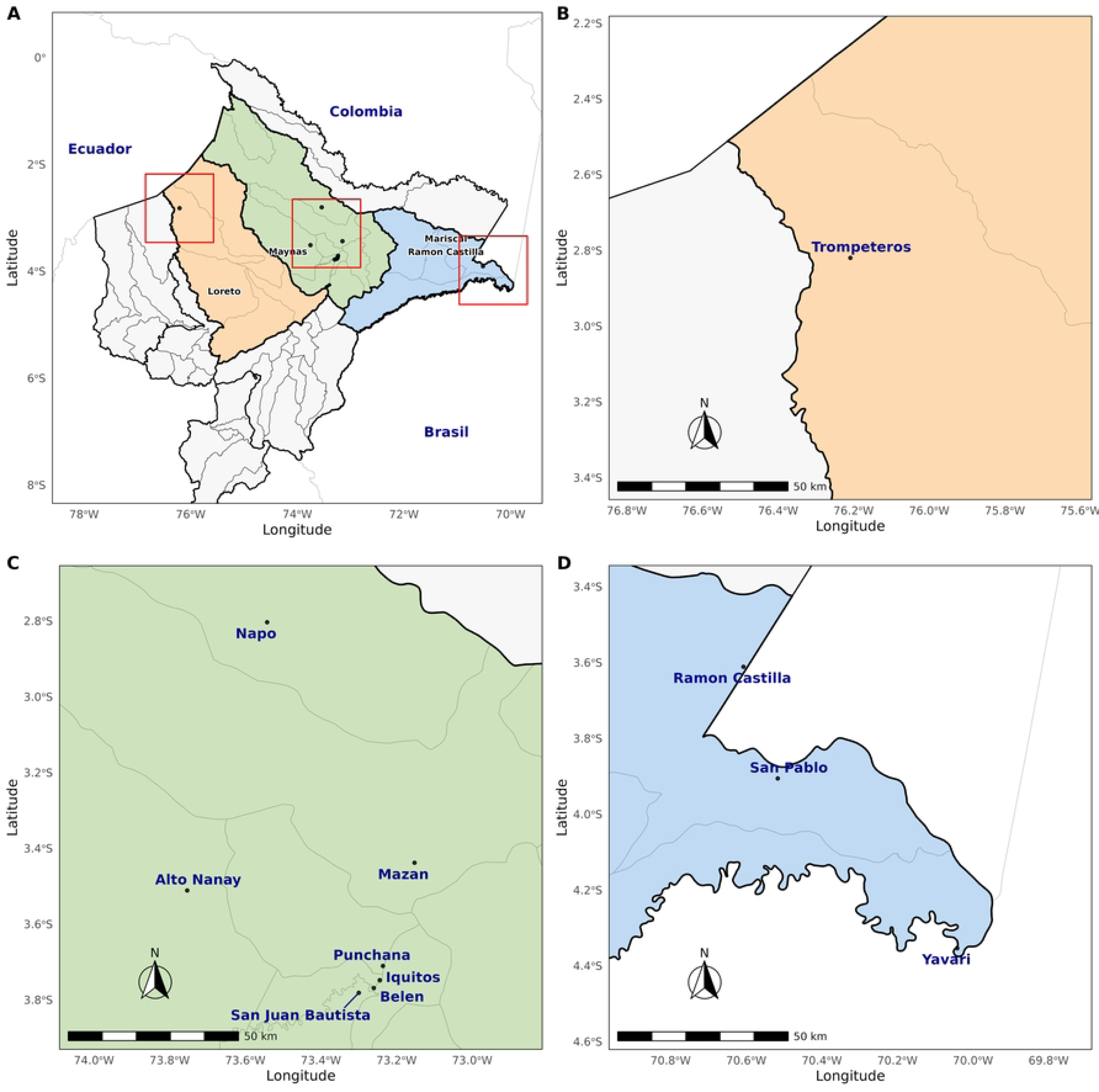
Map of included study sites in the Peruvian Amazon. A. Provinces in the Loreto region. Three provinces were included in this study (Loreto: orange, Maynas: green, Mariscal Ramon Castilla: blue). Areas within red squared are shown in B-D. B. Loreto province, depicting the Trompeteros district. The remote indigenous Nueva Jerusalen community is settled there. C. Maynas province, with many districts with peri-urban communities. D. Mariscal Ramon Castilla province, depicting 3 districts with border communities with Colombia and Brazil. Maps created an in-house script in R using the R-packages used R libraries SF, ggspatial, ggrepel.

The samples included here were collected in previously published studies from 2007-2008 (n=41) [31], 2016 and 2017 (n=54) [32], from population-based cross-sectional surveys in Mazan district in 2018 (n=13) [33, 34], and n=32 samples collected in 2014 by passive case detection (PCD) at the health center at San Juan district in Iquitos (S1 Table). In addition, samples from Ramon Castilla, Yavari and Trompeteros districts were collected in collaboration with MINSA authorities as part of surveillance activities in remote border communities in the Loreto region. Collections in Ramon Castilla (n=2) and Yavari (n=20) were conducted in December 2018 through active case detection (ACD) and PCD, respectively. Samples from Trompeteros (n=68) were collected during 3 weeks at the end of November as part of ACD visits and during April - May 2020 by PCD (Fig 2 and S1 Table).

**Figure 2.**
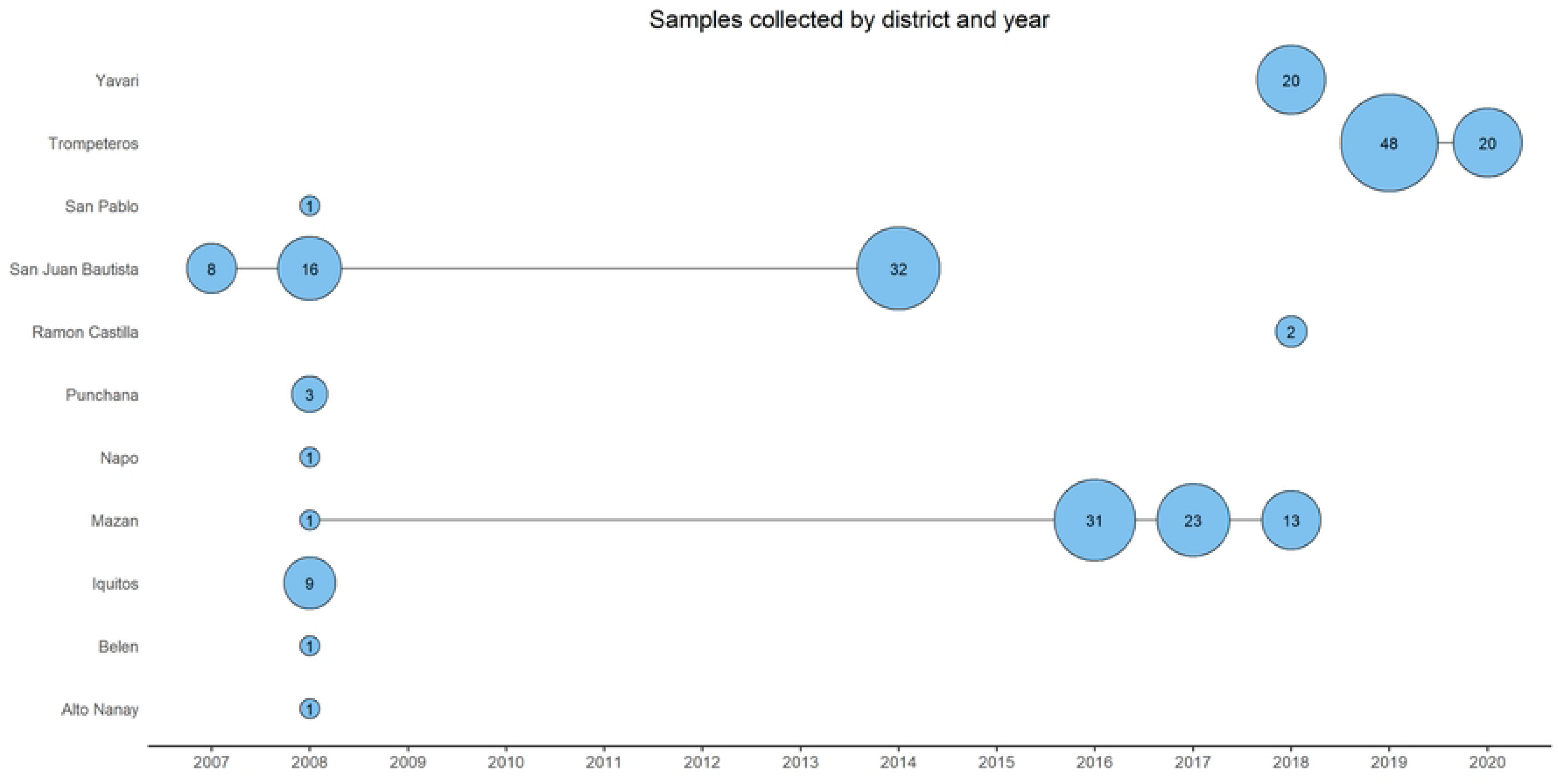
**Overview of samples from Peru over time (year) and district**.

### Ethical considerations

Samples from previous studies with consent for future use for malaria research were studies with protocols registered in the Decentralized System of Information and Follow-up to Research (SIDISI) of the University Directorate of Research, Science and Technology at Universidad Peruana Cayetano Heredia (UPCH), to be then evaluated by UPCH Institutional Research Ethics Committee (CIEI) prior to its execution (SIDISI codes 52707, 61703, 66235, 101518, 102725). In Yavari, Ramon Castilla and Trompeteros districts, samples were collected as part of routine surveillance activities by MINSA. MINSA authorities transferred these samples to the UPCH team for research activities (SIDISI project 102725). Secondary use of all samples for the purpose of genomic surveillance of malaria was approved through the Institutional Review Board of the Institute of Tropical Medicine Antwerp (reference 1417/20).

### DNA extractions and quantification

DNA from all samples was extracted using the E.Z.N.A Blood DNA Mini Kit (Omega Bio-tek, Georgia, USA) or QIAmp DNA Blood Mini Kit (Qiagen, Germany) following the manufacturer’s instructions. For DNA extraction using the E.Z.N.A kit, we used 40uL of whole blood, or a ∼0.72 cm^2^ piece of dried blood spot (DBS); DNA was eluted in a final volume of 50 μL. For DNA extraction using the QIAmp kit, we used two punches of DNA (0.6 cm diameter); DNA was eluted in a final volume of 100μL. *P. vivax* parasitaemia was quantified by 18S rRNA qPCR [35] and/or Pv mtCOX1 qPCR [36].

### AmpliSeq library preparation and bioinformatics

Pv AmpliSeq library preparation was performed using AmpliSeq Library PLUS for Illumina kit (Illumina), AmpliSeq Custom Panel design (*i.e.* the primer pools as per the new design; S2 Table) and AmpliSeq CD Indexes (Illumina) as per the manufacturer’s instructions. Library preparation was performed on 7 µL of undiluted DNA as previously described [23], following similar steps as the protocol for *P. falciparum* AmpliSeq [37]. Briefly, target regions were amplified in two reactions, and then combined for final library preparation. Libraries were quantified using Qubit v3 High sensitivity DNA kit (Invitrogen, Massachusetts, USA), and subsequently diluted to 2nM with low Tris-EDTA buffer, and pooled. Denatured library pool (7 pM) was loaded on a MiSeq system (Illumina) for 2x300 paired-end sequencing (Miseq Reagent Kit v3, Illumina) with 1-5% PhiX spike-in (Illumina).

FASTQ files were processed with an in-house analysis pipeline on a Unix operating system computer, as previously described [23], generating a jointly-called VCF file with variants (SNPs and INDELs) detected in the targeted regions (scripts are available at https://github.com/Ekattenberg/Plasmodium-AmpliSeq-Pipeline). Variants were hard filtered (QUAL>30, overall DP>100, MQ>50, QD>1.0, SOR<4, GT depth >5) and annotated with SnpEff (v4.3T) [38], resulting in 3862 high quality genotypes (incl. all variant types, *e.g.* SNPs and indels). Per locus filtered depth of coverage (format field DP in the vcf) was used to calculate the median depth of all loci per sample or per amplicon. Aligned coverage was calculated as the number of bases passed filter divided by the number of bases (59815 bp) targeted in the Pv AmpliSeq v2 Peru.

For the analysis of genomic surveillance use cases in Peru, we included 230 samples (Fig 1) with good quality data (<25% missing genotype calls for all variants, mean coverage >15), and retained only one library of replicates (with lowest missingness).

### Data analysis

Genetic diversity, expressed as expected heterozygosity (*He*), was calculated using polymorphic barcode (40/41 SNPs) genotypes from the vcf with the adegenet package in R [39, 40]. Nucleotide diversity (*pi*) was determined by sliding across the target regions in the genome in 500-bp windows using vcftools and a vcf file with all biallelic SNPs detected by the assay. DAPC with cross-validation was performed to infer population structure based on haplotypes across districts and years [30]. Associated allele loadings for the first four components in the DAPC were determined.

Genetic differentiation, expressed as fixation index (F_ST_), was calculated with the R package hierfstat [41] using all biallelic SNPs detected by the assay. Within-host infection complexity was assessed using within-sample F-statistic (Fws) with the R package moimix [42] using all biallelic SNPs detected by the assay. Fws ≥ 0.95 was considered a proxy for a monoclonal infection.

We created a list of variants of interest (S3 Table) that included variants in genes reported in the literature as potentially associated with *P. vivax* antimalarial resistance. The list was supplemented with non-synonymous variants detected in the target genes that contributed to the variation in the DAPC. Haplotypes were created by combining genotypes of variants of interest.

To measure pairwise identity-by-descent (IBD) between samples, PED and MAP file formats were generated using VCFtools. The level of IBD-sharing was calculated using the isoRelate package in R [43] with settings as previously described [23]. Network plots (at thresholds of 99%, 50% and 10% IBD) were created using the igraph package in R to visualize the relatedness between samples (95% and 50%) and connectivity between districts (10%).

A likelihood-based classifier was used to predict the origin of *P. vivax* isolates using the 33-SNP vivaxGEN-geo barcode, using the vivaxgen geo framework (https://geo.vivaxgen.org/) and reference dataset [44].

## Results

### Barcode and assay performance

We designed a 41-SNP Pv Peru Barcode, with in-country resolution (*i.e.* able to separate between distinct *P. vivax* populations in Peru based on variability of allele frequencies between districts and provinces) using *P. vivax* genomes from Peru (n=130) [17]. SNPs in the barcode were selected based on their contribution to geographically distinct genetic clusters in the DAPC. Allele frequencies (AF) of the 41 SNPs in the barcode were evaluated in all *P. vivax* genomes n=1474) originating from 31 countries from the same genomic dataset. Minor allele frequencies (MAF) of all SNPs varied between 0.01-0.49, with a median of 0.16 (S4 Table). Within genomes from Peru, most SNPs (63%) had a MAF>10%, with 10 alleles observed at MAF<5%. The MAF in isolates from multiple countries in Latin America (including Mexico, Panama, Colombia, Brazil) were similar (median MAF 0.09, range 0.01-0.49) to the MAF found in Peru. In addition, the Pv Peru Barcode was capable of separately clustering the Latin American population in PCA analysis, with increased resolution using n= 753 loci (*i.e.* all biallelic variants detected in Peruvian isolates with the Pv AmpliSeq v2 Peru), separating the samples from different countries with distinct subpopulations (Fig 3), matching patterns previously observed in the whole genome dataset. Altogether, this demonstrates a wider applicability of the Pv AmpliSeq v2 Peru assay to the Latin American region and the optimal resolution of the Pv Peru barcode to differentiate parasite populations by country in Latin America.

**Figure 3.**
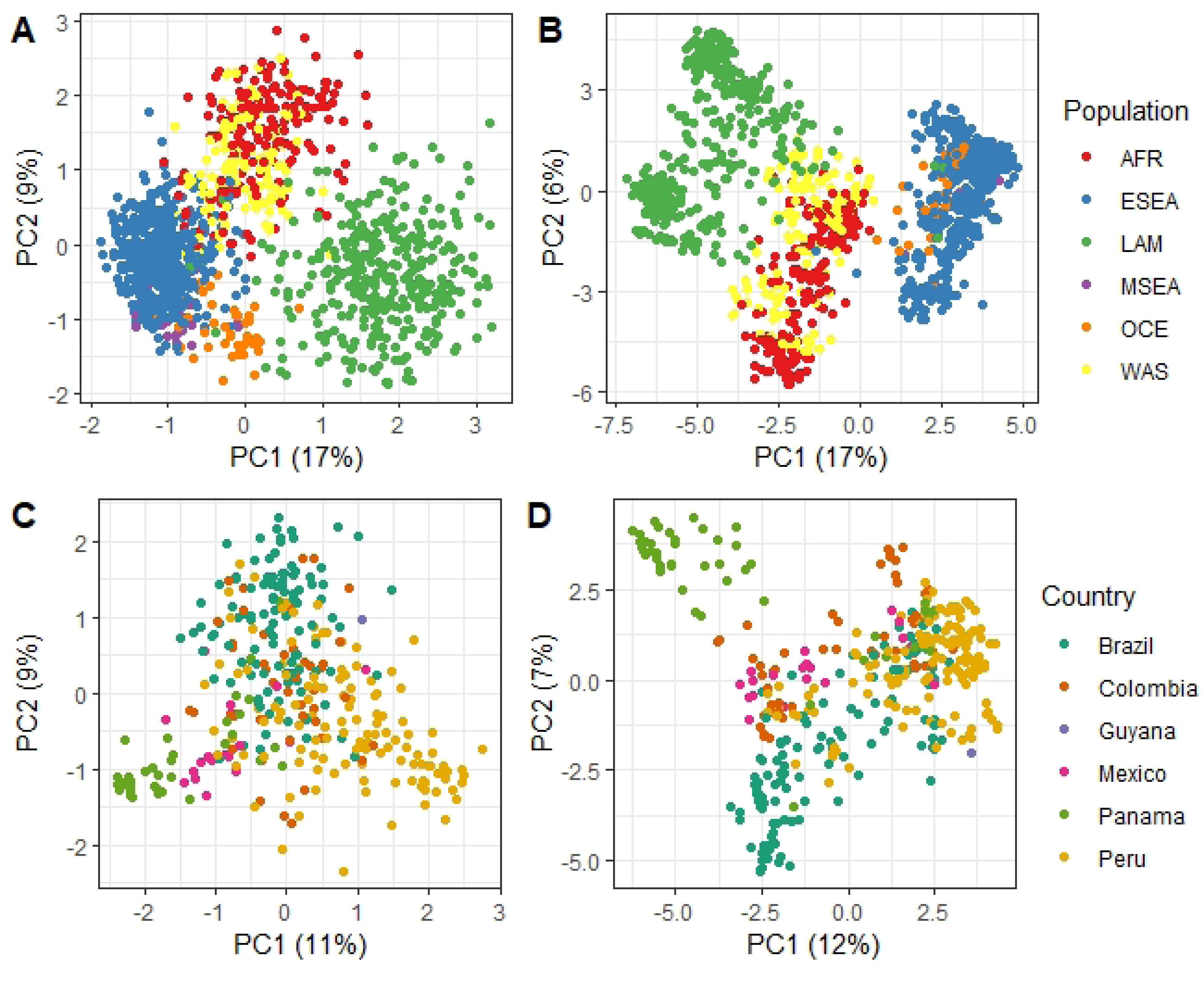
Principal Component Analysis of *P. vivax* genomes dataset. (23), filtered on the SNPs in the Pv Peru Barcode only (A & C), and all biallelic SNPs detected by the Pv AmpliSeq v2 Peru assay in the Peruvian Amazon samples (B&D). The samples (dots) are colored according to the originating population (regions or country).

The Pv Peru Barcode was experimentally validated with the Pv AmpliSeq v2 Peru assay using *P. vivax* isolates (n = 230) from several regions in Peru (Figs 1 and 2). All 41 targeted SNPs in the barcode were amplified successfully, with a median MAF of 0.24 (range: 0.01-0.49, mean ± standard deviation: 0.25 ± 0.14). For the SNP PvP01_13_v1_32509, only the reference allele was detected. We identified for each of the barcode amplicons how many biallelic SNPs were detected in the samples (median 14, range 4-28), which could potentially be used as microhaplotypes in a haplotype-based approach (S4 Table). The Pv AmpliSeq v2 Peru assay generated a high number of reads per sample (median 119033, interquartile range [IQR] 59535-243132 reads/sample after trimming low-quality reads), with a median of 99.4% (IQR 97.7-99.9%) of reads aligning in pairs to the PvP01 reference genome. The median depth of coverage of aligned high-quality reads past the filter (DP) was 335 reads (IQR 121 to 859) (S1 Fig). There were 4 (1.8%) amplicons with low mean DP-values (<10) (pvama1_4, pfdhfr_8, pvmdr1_1, and PvP01_14_v1_2841138), and 1 amplicon (0.44%) had high mean DP-values (>200) (pvcrt_11).

### Spatial-temporal patterns in transmission intensity

Complexity of infection (COI) and genetic diversity – expressed as expected heterozygosity (*He)* and nucleotide diversity *(pi)* – were used as a proxy of transmission intensity. Both parameters were compared between parasite isolates from different districts and years (Fig 3). *He* was estimated using the SNPs-barcode positions only, and *pi* was measured over all biallelic variants detected in the samples. Moderate levels of *He* were found in all districts, except in Yavari, which had a significantly lower diversity (mean *He* = 0.0121; p < 0.0001) (Fig 4A). *Pi* was significantly higher in Iquitos compared to its surrounding districts Mazan and San Juan Bautista (adjusted p<0.0001). In addition, similar to the observed patterns in *He*, *pi* was significantly lower in Yavari (adjusted p<0.0001) compared to all other districts (Fig 4C). *Pi* was higher in 2007-2008 than in later years (adjusted p<0.001, Figure 4D), alongside a period of intensification of malaria control and reduction of cases in Peru. No temporal trends were observed in *He* (Fig 4B). Diversity in samples from 2018 was lower than other years, however this is likely a result of the low diversity in Yavari in 2018, which made up the majority of samples in this year. In addition, we found polyclonal infections in 5 districts, all of them at a low proportion (10 - 35.3%) (S2 Fig).

**Figure 4.**
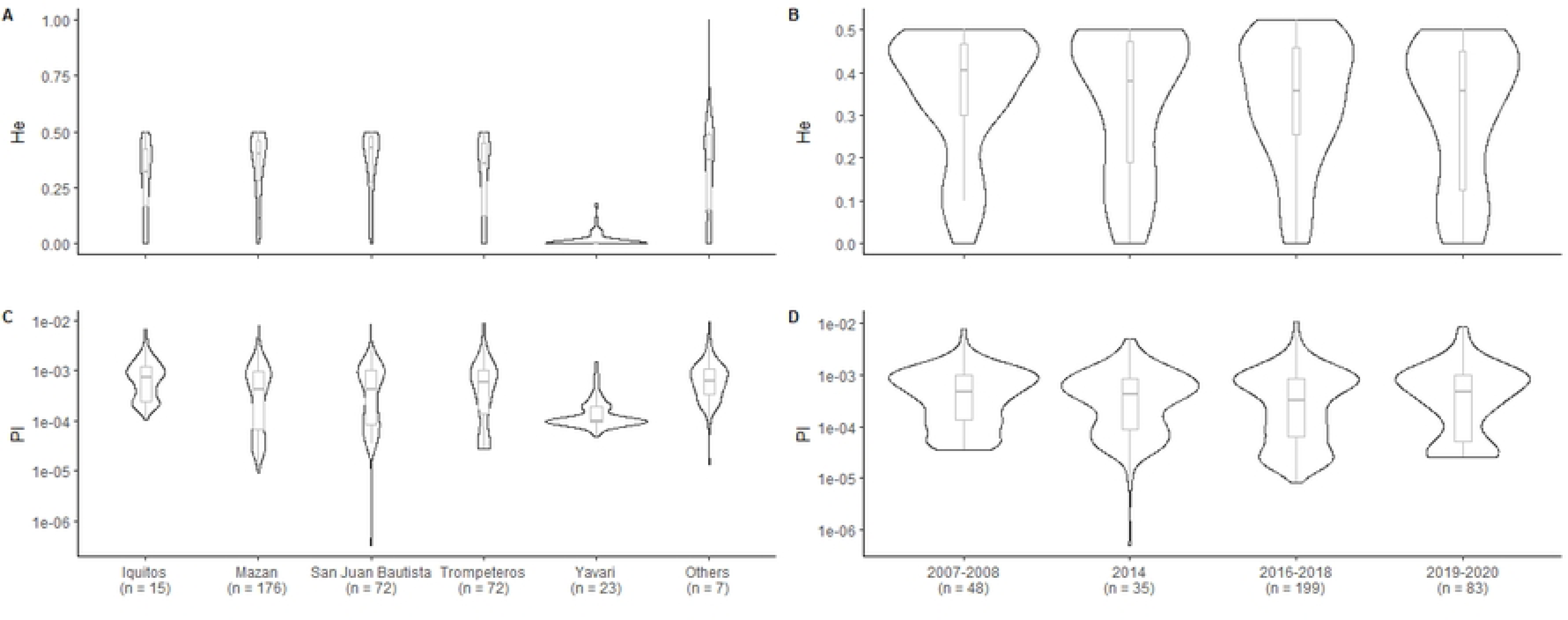
Molecular markers for transmission intensity. Violin plot of Expected heterozygosity (*He*) of 40-SNP barcode positions (A & B), and nucleotide diversity (*pi*) (C and D) measured across the targeted region in 5000 bp windows by district (A& C) and years (B & D).

### Population structure and connectivity

The geographic differentiation and structure of parasite populations was investigated at the district level. Estimation of pairwise genetic differentiation (Fst) using the biallelic SNPs detected showed little differentiation (Fst: 0.02 - 0.07) between districts; again, except Yavari, which was highly differentiated (Fst: 0.39 - 0.55) from all other districts and in particular from Iquitos (Fig 5A).

**Figure 5.**
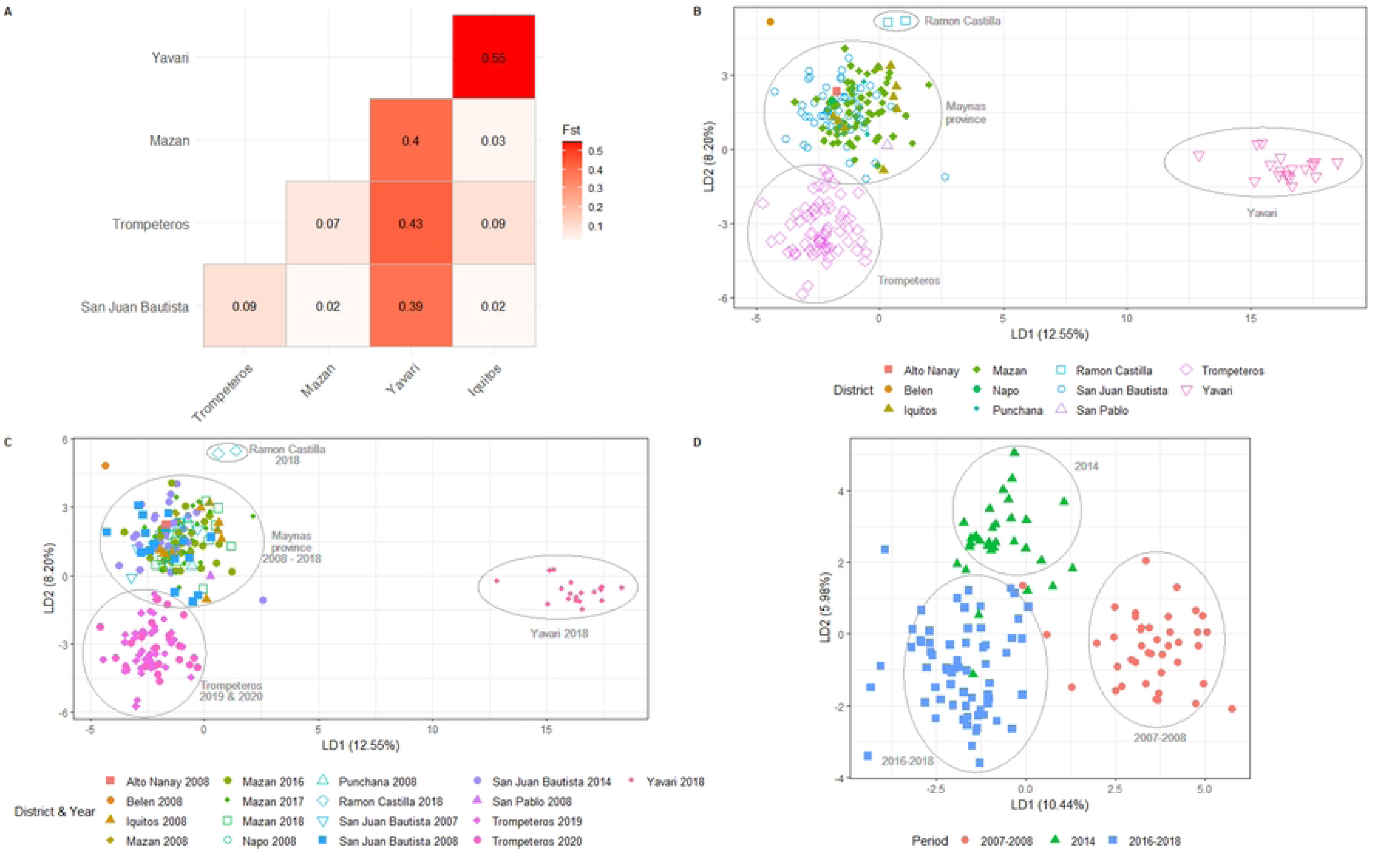
Population structure and genetic differentiation. Scatter plot of discriminant analysis eigenvalues 1 and 2 (LD1 and LD2) using all variants detected by the Pv AmpliSeq v2 Peru assay of Peru samples (n = 230) grouped by district (A) or district and year (B), and Maynas samples (n = 124) grouped by period (C). Heatmap of pairwise F_ST_ values between 5 districts in the Peruvian Amazon estimated with the hierfstat package in R (D).

We explored the population structure using DAPC and all variants detected with the AmpliSeq assay (N=3862). The first two components (PC) separate the communities in Maynas province from remote communities in Loreto and Mariscal Ramon Castilla provinces: Yavari (PC1), Trompeteros (PC2) and Ramon Castilla (PC2) (Fig 5B). This observed structure was not affected by sampling in different years, as clusters from different time points, but same areas, overlap in the DAPC (Fig 5C). However, within Maynas province, a temporal differentiation was found separating the samples in 3 clusters: 2007-08, 2014 and 2016-18 (Fig 5D).

From the variants contributing most to the first two axes of the DAPC analysis with all samples (Fig 5B), 10 out of 38 highest contributing alleles were SNPs in the barcode amplicons. Similarly, 12 out of 30 highest contributing alleles in the Maynas DAPC analysis (Fig 5D) were Pv Peru Barcode positions or other SNPs on the barcode amplicons (S5 Table), highlighting the contribution of these genomic regions to the population structure with sufficient resolution for within-country analysis in Peru. Highly-contributing alleles in genes of drug resistance interest, especially *pvmdr2* and upstream variants of *pvcrt*, were found, including both missense mutations, potentially under drug-induced selection pressure, as well as synonymous mutations, which do not impact the phenotype but reflect the population genetic background (S5 Table).

We assessed the connectivity of parasites in the Peruvian Amazon across space and time by measuring the genetic relatedness between isolates. We analyzed the pairwise IBD between samples within and between districts and years (Fig 6 and S3 Fig). High relatedness was seen mainly between samples from the same district, with a clonal population (sequences sharing IBD > 99%) in Yavari (Fig 6A). At moderate levels of IBD-sharing (50%), there was a lot of relatedness within Trompeteros samples (Fig 6B). At even lower levels of IBD (10%), samples from Yavari, Trompeteros, Mazan and San Juan Bautista become connected (*i.e.* high amount of sample pairs with at least 10% IBD), regardless of their geographical distance within these districts (Fig 6C). Many clusters of high IBD (50% or 90%) were found within one year, however, some sample pairs from different years with high relatedness were observed (S3 Fig).

**Figure 6.**
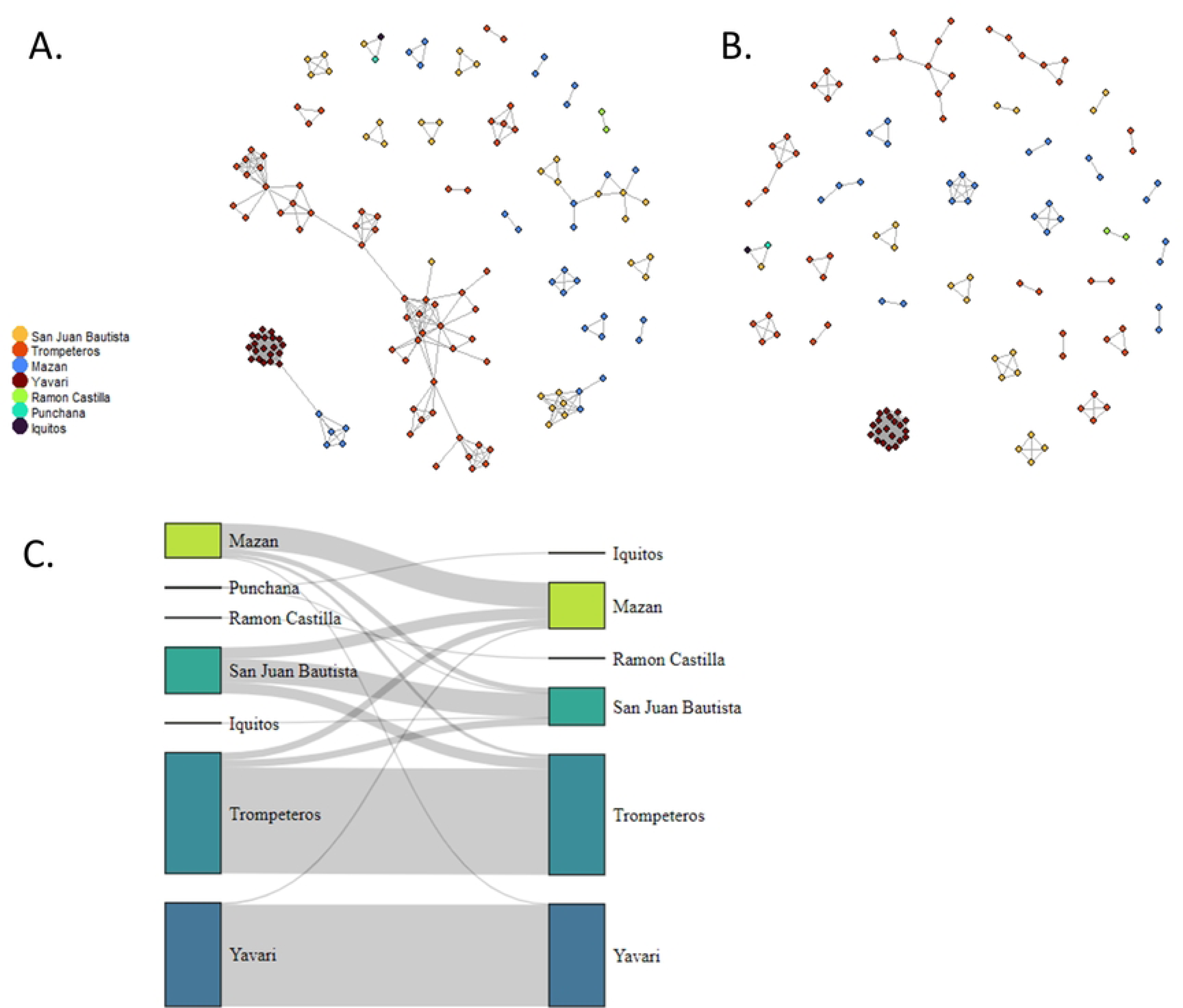
Parasite relatedness. Network of individual relatedness at (A) intermediate levels of relatedness (50% IBD threshold) and (B) Very high levels of relatedness, indicating clonal infections (95% IBD threshold) colored by district. (C) Network of connectivity between districts using low levels of relatedness (10% IBD threshold).

### Variants of Interest

We investigated the haplotypes in the genes that have a potential association with drug resistance in *P. vivax* and orthologue species as previously described [23]. Haplotypes were constructed of variants of interest (S6 Table), including variants with a potential association with resistance and non-synonymous variants contributing to the DAPC in Fig. 5A.

High variability in the genes *pvmdr1, pvdhps, pvdhfr, pvmdr2, pvp13K, pvmrp1,* and *pvmrp2* was found in most districts (S4 Fig), whereas low variability was observed in *pvcrt* (only one sample with an amino acid change at position 275, F>V) and *pvK13* (only one haplotype). In *pvdhps*, we observed the sulphadoxine-resistance associated mutation A383G in 61.7% (142/230) of samples, alongside other phenotypically uncharacterized mutations (S6 Table). In *pvdhfr*, we observed the pyrimethamine-resistance associated mutations S58R (125/230, 54.3%) and S117N (3/230, 1.3%), alongside other uncharacterized mutations, including S58K (S6 Table). Two observed different nucleotide changes (codons AGA and AGG) resulted in the S58R amino acid change in *pvdhfr*. The A553G mutation in *pvdhps* and mutations F57L, T61M, S117T in *pvdhfr* were absent from the samples from Peru. In *pvmdr1*, we observed the CQ resistance-associated mutations Y967F (CQ and AQ) and F1076L (CQ) in 15.2% (35/230) of samples in Peru, always combined in the same haplotype that also carried the S698G mutation (S6 Table).

The DAPC analysis identified differences between populations (Figs 5A and 5B), including mutations in *pvdmt2* (S277Y), *pvmdr2* (G305S, S1038N, D1447E, A1450V, and T1480A), *pvmrp1* (H1586Y, I1478V, G1419A and K36Q), *pvmpr2* (N1251Y, P330S), and large indels in *pvdhfr* (at amino acid positions 618 and 639) and *pvp13k* (indels at amino acid positions 86, 106-108, 834, and mutations E1456K and E810G), but the phenotypes of these variants have not been characterized. Yaravi samples presented haplotypes in most of the genes (*pvmdr1, pvdhps, pvdhfr, pvmdr2, pvp13K, pvmrp1,* and *pvmrp2)* that were found exclusively in this location or were rare in other areas.

We also investigated the variability in the antigenic *pvama1* gene, with 6 observed haplotypes of 6 non-synonymous variants detected in the amplicons targeting the highly variable regions in this gene (S6 Table). Moderate variability was found with all haplotypes detected in all years (S5 Fig). A high proportion of samples with Haplotype 19 was observed in 2007-08 (39%), which decreased in later years (16% in 2014, 24% in 2016-2018 and 4% in 2019-2020). In contrast, haplotype 4 was predominant in 2016-2018 (24%), mainly corresponding to isolates from border districts Yavari and Ramon Castilla (S5 Fig).

### Predicting the origin of infections

We predicted the origin of samples from different districts in the Loreto region using the SNP vivaxGEN-geo barcode included in the panel. As expected, most samples from Mazan, San Juan Bautista, Iquitos and Trompeteros were predicted to originate from Peru (predicted value ranging between 50 – 75%), with a small proportion of samples with principal predictions to originate from Brazil (predicted value ranging between 22 – 35%) and Colombia (predicted value ranging between 3 – 25%) (Fig 7). Samples from Yavari, Ramon Castilla and San Pablo - all border communities - were more similar to Brazilian isolates (Fig 7).

**Figure 7.**
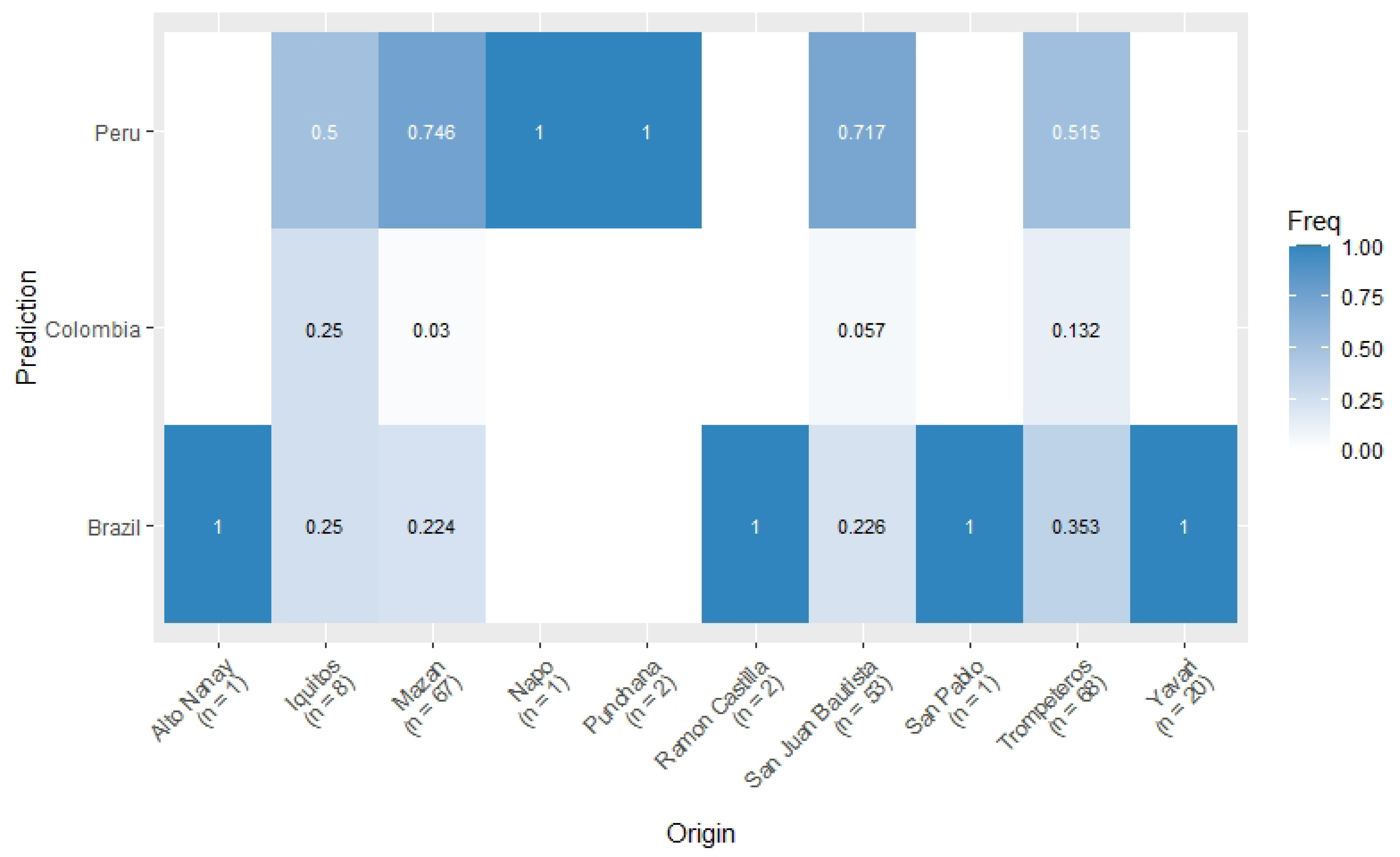
Prediction of origin of *P.vivax*. Heatmap of frequency of the prediction of origin in the vivaxGEN-geo likelihood classifier (y-axis) by sample collection site (x-axis).

## Discussion

In this study, we designed a Pv Peru Barcode capable in combination with the Pv AmpliSeq panel [23] to differentiate parasite populations from not only Peru, but the wider region of Latin America at a high resolution. We successfully applied our Pv AmpliSeq v2 Peru assay to a retrospective sample collection (n=230) from 11 districts in the Peruvian Amazon, and observed marked differences between peri-urban, remote and border communities in Peru, with overall a heterogenous *P. vivax* population with high diversity in the Loreto region, but moderate to low diversity at the community level. This pattern of low-local but relatively high-regional genetic diversity has been described previously using microsatellites [45] and is likely caused by the effect of random genetic drift in small populations that remain relatively isolated from each other, where rare alleles disappear (decreased local diversity translated in a reduction of He, and high relatedness IBD), while differentiation between sites increases. In contrast, in peri-urban communities close to Iquitos, the regional capital, a relatively high genetic diversity is observed with connectivity within and between districts, and a temporal drift of the parasite population, in concordance with previous reports [19, 46].

The observed connectivity between the peri-urban settings in Maynas province is likely due to human economic activities and travel to the economic centers in the areas [47]. The presence of malaria corridors in the Peruvian Amazon due to human mobility and other socioeconomic factors has been previously reported [7, 21]. The effects of human movement on malaria transmission in peri-urban areas of the Peruvian Amazon have been well-studied, though less is known about how migration and mobility affect the spread of malaria to and from remote indigenous and border communities.

In the remote isolated area with an indigenous population (Trompeteros district in Loreto province), a genetically distinct parasite population was observed with moderate diversity and high relatedness between isolates in the community and limited connectivity to districts in Loreto province. While in the remote community in the Yavari district (Mariscal Ramon Castilla province), along the border with Brazil, a highly differentiated clonal *P. vivax* population was observed, with distinct haplotypes in drug resistant genes and *pvama1*. This population was predicted to be more similar to Brazilian isolates, and indeed shows little connectivity with the populations in the other provinces in Peru. This highly differentiated clonal population could be a possible outbreak introduced from Brazil. Out of more than 160 collected samples from symptomatic patients in that community, 22% (36/163) tested positive for *P. vivax* through PCR analysis. However, due to limited epidemiological data at that time, an outbreak cannot be officially confirmed. Besides typically clonal reported *P. falciparum* outbreaks [48–50], few *P. vivax* outbreaks have been previously reported in Peru [32].

In addition, due to the vicinity of Yavari to the Brazilian border it is also possible that the parasite population in this region shares ancestry with populations in Brazil, which could result in a similar genetic pattern, and the parasite strain was not recently imported. Predictions of origin of parasites in this region is also limited by the low number of South American isolates in the reference dataset in the applied tool, which could be improved by including more recent genomes from this region from other studies [16, 17]. In addition, there were 7 (3%) samples from Peru with predictions from Vietnam, Afghanistan and Iran. While it is possible that cases were imported from areas outside of South America, it is more likely that this is a result of incorrect predictions due to missing data in the barcode used for the prediction (3 samples had 18 – 30% missingness in the global barcode), or inaccuracies in the reference dataset (proportion missingness of all variants: 4 – 28%), as this was also observed in our earlier study [23].

The highly related parasites circulating in these remote districts provide evidence of local isolated transmission in these hard-to-reach communities (many hours or days of riverine and road travel from Iquitos). Personal communication with inhabitants in these areas during field work indicates some mobilization to nearby communities for work or social activities, but not to further away areas such as Maynas. However, we described the relatedness between *P. vivax* parasites found in these remote communities with the parasite populations closer to the economic center. Future studies focused on human mobility patterns and related socio-epidemiological features in hard-to-reach communities are needed to better understand the factors driving malaria transmission in these communities.

These differences in gene flow and transmission dynamics resulting in distinct genetic patterns are important for the NMEP and their strategy for malaria control and elimination. First, it’s important to inform implementation of local *vs.* regional interventions in order to ensure all areas progress towards malaria elimination. Second, the gene flow between the Yavari population with a neighboring country, but also with isolates from other districts, shows the importance of increasing surveillance in border areas (not only with Brazil, but also Colombia and Ecuador) and creating connections with health authorities from these countries to collectively better control malaria [51]. In addition, stable local transmission in hard-to-reach indigenous communities such as in Trompeteros can be targeted separately. There is a lower risk of malaria returning after elimination in these areas due to geographical distance and cultural differences. However, for the same reasons, MINSA interventions do not usually reach them or only on a temporary basis. In summary, district or community-level approaches with targeted interventions, as previously proposed [52], may be useful in Peru given its heterogeneous context.

Other regions in Peru, such Amazonas region (next to Loreto) or the Northern Coast, were not included in this initial study and more work is needed to fully characterize the full *P. vivax* population in Peru, especially considering the observed low local, but high regional diversity. However, compared to other previous studies, we included more districts and remote areas spanning a larger period [19–21]. To assess the full diversity of Peruvian parasites, a more systematic sampling approach, possibly linked with NMCP and/or NMEP activities is needed. Collection of samples should be performed regularly through multiple sentinel sites across the country, including hard-to-reach communities and potentially using current active case detection strategies when number of cases increases over a defined threshold at the community level.

Distinct haplotypes in the genes putatively associated with drug resistance were detected in Peruvian parasites. However, in contrast to *P. falciparum,* no validated markers of resistance are known for *P. vivax*, except for SP-resistance [53]. Therefore, the observed haplotypes are not characterized for CQ or ART resistance in *P. vivax*, which are the first-line drugs for *P. vivax* and *P. falciparum* malaria in Peru, making it difficult to interpret these results considering the national treatment guidelines. In addition, while the WHO recommends routinely monitoring antimalarial efficacy every 2 years [54], the latest treatment efficacy studies (TES) for *P. vivax* in Peru were conducted about 10 years ago [55, 56]. *Ex-vivo* assays are another approach the characterize resistance profiles of parasites, which need to be conducted quickly after sample collection, and require laboratory facilities in the vicinity of sample collection sites [57]. Reports from Porto Velho and Mâncio Lima in Brazil conducted in 2012 - 2015, for example, indicated that there was no chloroquine resistance at that time [58, 59]. Another study with Iquitos samples from 2015-2019 in Peru reported 3.3% (1/30) of isolates with chloroquine resistance [60]. However, these assays are not suitable for low parasitaemia infections that are common for *P. vivax*, complicating routine analysis [57].

The Pv AmpliSeq assay has been successfully applied here to low density samples from Peru, using the previously determined threshold of 5 parasites per microliter [22]. Accurate quantification through standardized qPCR protocols and utilizing Ct-value cut-offs, which better represent parasite DNA quantity than difficult to standardize diluted samples with known parasite densities, can best ensure precise sample selection for efficient surveillance protocols. In addition, maintaining DNA integrity in DBS samples, which are ideal for sample collections in remote communities due to easy transport to reference laboratories [61], is important to ensure good performance of the assay.

In summary, we demonstrated the heterogeneity of *P. vivax* transmission in the Peruvian Amazon in different settings by applying the Pv AmpliSeq v2 Peru assay. Results show local low diversity and connectivity patterns that can be important for the NMEP to improve and adapt strategies depending on the local epidemiology, ecology, and human population. Genomic surveillance of malaria using the Pv AmpliSeq v2 Peru can be a useful tool when applied routinely in the country and has the potential to be used in other countries in South America with similar allele frequencies of the barcode SNPs. To ensure sustained progress in malaria control and achieve long-term elimination, it’s essential to prioritize the effectiveness of NMCP/NMEPs by optimizing resource utilization and strategically implementing necessary changes. Informed decision-making, guided by relevant data and the increasing recognition of the potential of genetic surveillance tools, is crucial for addressing specific challenges related to *P. vivax* control and elimination.

## Data Availability

Raw data (FASTQ files) are available at the SRA under BioProject accession number PRJNA1055117 and individual sample accession numbers are listed in supplementary data file S1 Data. Data preparation scripts are available at https://github.com/Ekattenberg/Plasmodium-AmpliSeq-Pipeline. All other data are included in the manuscript and supporting information.

## Acknowledgments

First, we want to thank all the participants, field workers, and related staff of the different projects included in this work for making the sample collection possible and their material available for this study. We would also like to acknowledge Prof. Dr. Jean-Pierre Van geertruyden (Global Health Institute, UAntwerp) and Dr. Hugo Valdivia (Naval Medical Research Unit SOUTH) for their contribution on Trompeteros data and sample acquisition.

## Supporting information Captions

**S1 Table. Summary of samples and studies included in the Pv AmpliSeq assay.**

**S2 Table. Pv AmpliSeq v2 Peru design.** Primer sequences and target regions in the AmpliSeq design.

**S3 Table. Variants of interest list.**

**S4 Table. Allele frequencies of barcode positions and microhaplotypes.** Allele frequencies were determined in a WGS dataset and are reported for worldwide, Latin American countries only, and Peru.

**S5 Table. Variants contributing to the population differentiation in the DAPC.**

**S6 Table. Haplotypes and haplotype frequencies of the variants of interest observed in Peru and control samples.**

**S1 Fig. Distribution of depth of coverage for each amplicon in the Pv AmpliSeq v2 Peru assay.**

**S2 Fig. Proportion of polyclonal infections detected in each district.** Within-host infection complexity was used as a measure of complexity of infections, using within-sample F-statistic (Fws) ≥ 0.95 as proxy for a monoclonal infection.

**S3 Fig. Parasite relatedness.** Network of individual relatedness at (A) intermediate levels of relatedness (50% IBD threshold) and (B) Very high levels of relatedness, indicating clonal infections (95% IBD threshold) colored by years.

**S4 Fig. Parasite relatedness and haplotypes of variants of interest.** Network of individual relatedness at intermediate levels of relatedness (50% IBD threshold) colored by haplotypes for the different genes.

**S5 Fig. Distribution of *pvama1* haplotypes by year and district**.

**S1 Data. Database of samples**. List of samples and data included in the study.

